# Promoting HIV care continuum outcomes among people who use drugs and alcohol: A systematic review of randomized trials published from 2011 to 2021

**DOI:** 10.1101/2022.07.26.22278090

**Authors:** Eileen V. Pitpitan, John Mark Wiginton, Raul Bejarano Romero, Dania Abu Baker

## Abstract

Substance use remains a robust predictor of HIV infection, as well as a serious impediment to progress across the HIV care continuum for people living with HIV. As such, the careful design and implementation of interventions uniquely tailored to target substance use and HIV care behaviors remain paramount. A necessary step in these efforts is to understand the extent to which HIV care interventions have been efficacious in helping people who use substances progress across the HIV care continuum. Using PubMed and ProQuest databases, we performed a systematic review of randomized trials of HIV care continuum interventions among people who use substances published between 2011 and 2021, the treatment-as-prevention era. Existing systematic reviews and studies in which less than half of those sampled reported substance use were excluded. We identified ten studies (total N=5410; range: 210-1308), nine of which intentionally targeted substance-using populations. Four of these studies involved use of at least one of several substances, including alcohol, opioids, stimulants, and/or marijuana, among others; three involved injection drug use only; one involved methamphetamine use only; and one involved alcohol use only. One study targeted a population with incidental substance use, which involved use of alcohol, injection drug use, and non-injection drug use. Viral suppression was targeted in 8/10 studies, followed by uptake/initiation of antiretroviral therapy (ART; 6/10), ART adherence (6/10), retention to care (4/10), and linkage to care (3/10). For each outcome, intervention effects were found in roughly half of the studies in which a given outcome was assessed. Mediated (2/10) and moderated (2/10) effects were minimally examined. The diversity of substances used in and across studies, as well as other characteristics that varied across studies, prevented broad deductions or conclusions about the amenability of specific substances to intervention. Moreover, study quality was mixed due to varying attrition and assessment measures (self-report vs biological/clinical). More coordinated, comprehensive, and targeted efforts are needed to disentangle intervention effects on HIV care continuum outcomes among populations using diverse substances.

## Introduction

Substance use remains a robust predictor of HIV infection across resource-diverse contexts and settings (Allen, Myers, & Ray, 2015; Huff et al., 2022; Rumbwere Dube, Marshall, Ryan, & Omonijo, 2018; Shuper et al., 2010). Specifically, a recent systematic review found that injecting drugs, smoking crack-cocaine, and binge-drinking (as well as similar behaviors by one’s partner) predicted HIV infection among adults in high-income countries (Rumbwere Dube et al., 2018), while separate reviews similarly reported that injecting drugs and using stimulants (such as methamphetamines) contributed to HIV burden in lower-resource environments (El-Bassel, Shaw, Dasgupta, & Strathdee, 2014). Pathways linking substance use to HIV infection include direct routes, such as needle sharing among people who inject drugs, and indirect routes, through behavioral disinhibition (e.g., through heavy alcohol use or stimulant use) in the form of condomless sex (El-Bassel et al., 2014; Heath, Lanoye, & Maisto, 2012; Huff et al., 2022; Rehm, Probst, Shield, & Shuper, 2017; Sandfort et al., 2017; Vosburgh, Mansergh, Sullivan, & Purcell, 2012).

Among people living with HIV (PLHIV), substance use has also been shown to impede progress at multiple stages of the HIV care continuum, from late HIV diagnosis to treatment failure (Huff et al., 2022), accelerating HIV disease progression (Carrico, 2011). For example, engaging in heavy alcohol use – harmful or hazardous alcohol use, binge-drinking, or levels of drinking that are consistent with those seen in alcohol use disorders – has been shown to hamper uptake/initiation of antiretroviral therapy (ART), decrease ART adherence and CD4 cell count, increase viral load, and overall accelerate HIV disease symptom onset (Azar, Springer, Meyer, & Altice, 2010; Molina, Simon, Amedee, Welsh, & Ferguson, 2018; Rehm et al., 2017; Shuper et al., 2010). Likewise, a qualitative study involving people engaged in injection and non-injection drug use (marijuana, heroin, cocaine, methamphetamines) found that substance use prevented or delayed HIV testing and linkage to and retention in care, as well as limited ART adherence (Kuchinad et al., 2016). Other research has demonstrated that injection drug use and stimulant use negatively affect retention in care and ART adherence (even resulting in discontinuation) and increase viral load (Carrico et al., 2007; Colasanti, Stahl, Farber, Del Rio, & Armstrong, 2017; Gonzalez, Barinas, & O’Cleirigh, 2011; Hinkin et al., 2007; Robison et al., 2008). Notably, research has indicated that substance use itself (stimulant use in particular) may facilitate viral replication, thereby leading to higher viral load, regardless of whether or not one adheres to ART (Baum et al., 2009; Carrico, 2011).

Given the substantial role that substance use continues to play in HIV care continuum outcomes, the careful design and implementation of interventions that are uniquely tailored to target substance use (or the mechanisms through which they impact the HIV care continuum) and HIV care behaviors (such as engagement in care and adhering to ART) remain paramount. A necessary step in these efforts is to understand the extent to which HIV care interventions have been efficacious in helping people who use substances progress across the HIV care continuum. Further, it is imperative to note instances in which key factors, informed by behavior change theory (Fisher, Fisher, Amico, & Harman, 2006; Starace, Massa, Amico, & Fisher, 2006), have been examined as potential mediators (e.g., HIV treatment self-efficacy or ART adherence mediating the path to viral suppression) and moderators (e.g., by substance used, level/severity of substance used, mental health, age) in analyses of intervention outcomes, as this would increase understanding of the mechanisms through which interventions have operated to impact outcomes, as well as highlight which subgroups of individuals have been more or less effectively impacted by interventions.

The objectives of this paper were twofold: (1) to review published literature from January 2011 to September 2021 to examine the extent to which HIV prevention and HIV care continuum interventions have been efficacious for people who use drugs and/or alcohol, and (2) to explore the extent to which mediators and moderators have been tested as part of the outcome analyses for these interventions. We selected 2011 as the starting point because this was the beginning of the treatment as prevention (TasP) era, when test-and-treat strategies were beginning to be understood and implemented (Cohen et al., 2011; Granich et al., 2011; WHO, 2012). A quantitative synthesis was beyond the scope and aims of the paper.

## Methods

### Eligibility criteria

A study was eligible for inclusion in the systematic review if it met the following criteria: 1) focused on people at risk for or living with HIV; 2) evaluated the efficacy of a behavioral intervention; 3) included people who reported active or recent drug or alcohol use (at least half of the sample), or the study was specifically focused on a substance-using population; 4) examined HIV prevention or HIV care continuum outcomes (defined as any of the following: uptake of HIV pre-exposure prophylaxis [PrEP]; PrEP adherence; linkage to HIV care; retention in HIV care; ART uptake, use, or adherence; HIV viral load or viral suppression; or immune function [e.g., CD4 count]); 5) used a randomized controlled trial design; and 6) sampled at least 200 participants. The latter two criteria were added toward the end of the article selection process to restrict the review only to studies using the gold-standard design for evaluating intervention efficacy, to eliminate smaller pilot trials, and because moderation and mediation analyses require dividing the sample across different subgroups, which is difficult or impossible with a small sample size. We did not include systematic literature reviews.

### Literature search

In September 2021, we conducted electronic searches of articles indexed in Pubmed and ProQuest between 2011 to 2021. We restricted the literature to peer-reviewed studies published from 2011 onward, as this year marked the beginning of the Treatment as Prevention (TasP) era.

#### Search strategy

Our PubMed search using the terms (HIV intervention) AND (prep OR pre-exposure OR treatment OR care OR adherence OR viral) AND (drug OR substance) yielded 2912 articles published since 2011 (Figure 1). Our ProQuest search using the terms (HIV) AND (intervention) AND (treatment OR care OR adherence OR ART adherence OR viral OR pre-exposure OR PrEP) AND (drug OR substance OR misuse OR dependence OR addict*) yielded 2678 articles published since 2011. We exported all 5590 records to DistillerSR (Evidence Partners, Ottawa, Canada), an online systematic review automation tool. After removing duplicates, we used the same software to subject the remaining 4999 articles to screening based on the aforementioned criteria. As part of our review process, we identified 19 articles that were systematic reviews of the literature about HIV and substance use and therefore relevant to the topic of this review (Abebe Moges, Olubukola, Micheal, & Berhane, 2020; Ahmed et al., 2018; Arrivillaga, Martucci, Hoyos, & Arango, 2013; Bazzi et al., 2019; Casale, Carlqvist, & Cluver, 2019; Feelemyer, Des Jarlais, Arasteh, & Uusküla, 2015; Glick et al., 2020; Govindasamy, Ford, & Kranzer, 2012; Hudelson & Cluver, 2015; Larney et al., 2017; Locher, Messerli, Gaab, & Gerger, 2019; Low et al., 2016; Mistler, Copenhaver, & Shrestha, 2021; Nachega et al., 2012; Nguemo Djiometio et al., 2020; Paquette & Pollini, 2018; Perelman, Rosado, Ferro, & Aguiar, 2018; Uthman, Oladimeji, & Nduka, 2017; Velloza et al., 2020). We examined these articles and their reference lists and found no additional studies in the reference lists that met our inclusion criteria.

**Figure 1.**
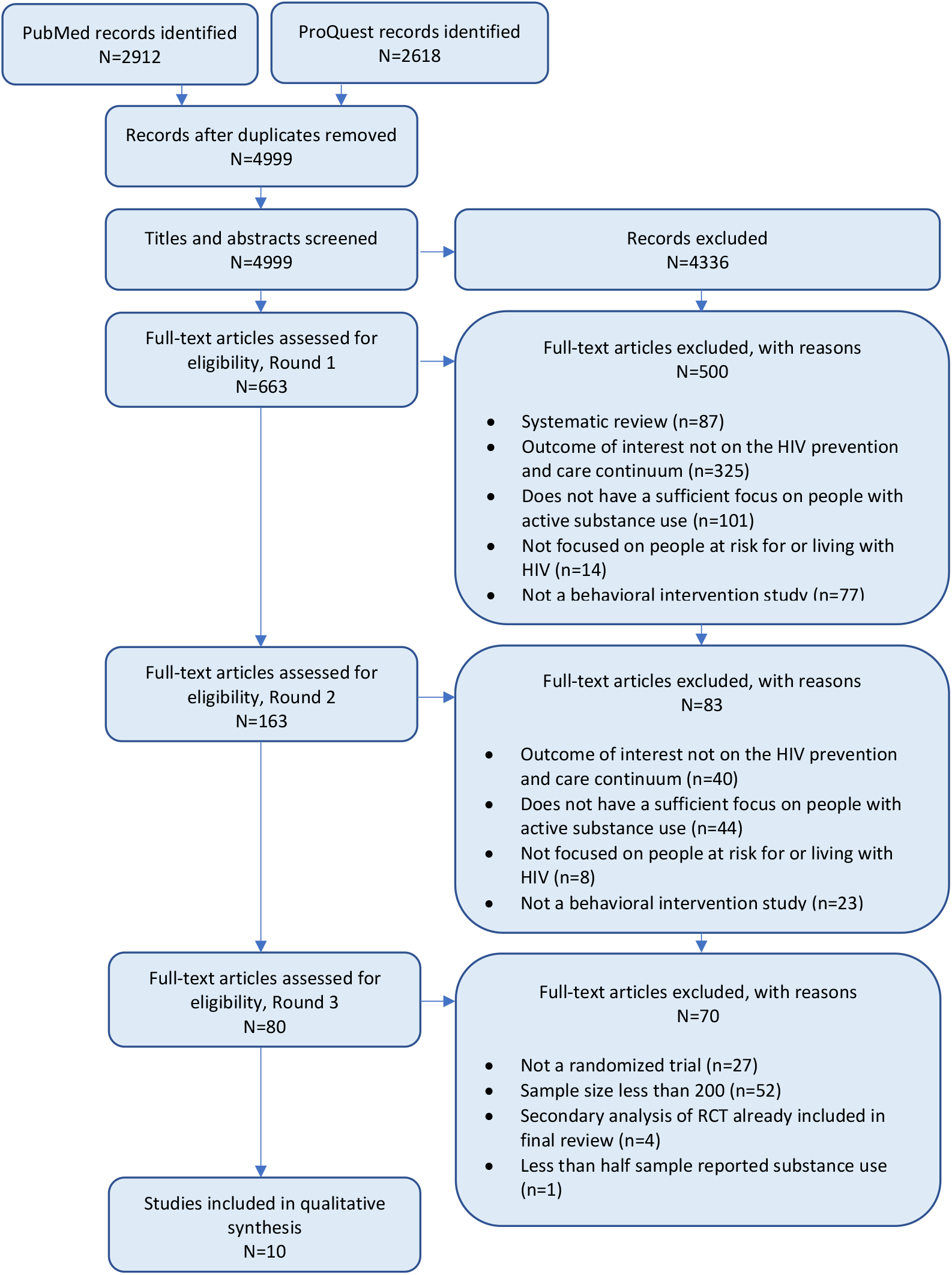
PRISMA flow diagram.

#### Selection process

Reviewers were not blind to the authors, funding, or any other characteristics of the studies reviewed. At the beginning of the screening and review process, two reviewers (DAB, RB) independently screened a small subset of the 4999 articles and discussed disagreements to promote rater reliability. The two reviewers then independently screened abstracts from the same subset of 1250 (25%) articles identified in the initial search to determine interrater reliability. Reviewers were instructed to include “HIV intervention studies” and to exclude editorials, opinion papers, and qualitative studies. Reviewers exercised liberal judgment for initial inclusion of potentially relevant articles for further review and met on a weekly basis to discuss and resolve disagreements. Interrater reliability across the two reviewers was high (kappa=0.99).

Subsequently, each reviewer independently screened 50% of the remaining abstracts, continuing with the liberal approach, which resulting in excluding 4336 articles and retaining 663 articles for full-text screening. For this process, reviewers independently screened the full-text of the remaining articles and adjudicated inclusion criteria to determine inclusion in the final sample. Reviewers considered the following questions to determine eligibility: 1) Is this a systematic review? 2) Is this study focused on people at risk for or living with HIV? 3) Does this paper include a focus on people with active substance use, either as described by the authors or as determined by the reviewer from the sample description? 4) Is this an intervention study? 5) Is the outcome of interest a variable in the HIV prevention and/or care continuum? Reviewers could indicate “yes” or “no” in response to these questions, and could also indicate “unsure” for questions 2 through 5. Any response other than “no” (indicating exclusion) for a given article resulted in the article’s being included in a second round of review to assess eligibility.

After this first round of full-text screening, 500 articles were excluded, with 163 articles being retained for further review. A second round of review resulted in 83 articles being excluded, leaving 80 articles for the next, final phase of review. It was at this third round of the article selection phase that we added the two additional inclusion criteria described previously: randomized controlled trial design and a sample size of over 200 participants. After this stage, 65 articles were excluded. An additional five papers were later excluded – four because they were found to be secondary analyses of a study already included in the final review, and one because less than half of the sample reported substance use (and substance use was imprecisely measured), leaving ten articles for the present review. Studies that were included in the final sample were agreed upon by the two reviewers and the lead author.

#### Data collection process

The two reviewers (DAB, RB) each examined five articles and extracted data from multiple domains: study authors; year published; recruitment time frame; population targeted (as described by the original authors); city or region where the study was conducted; sample size; mean or median age of the sample; name of the intervention being tested (if available); whether and how the intervention was described as being individual, community, structural, or multilevel; all intervention strategies (active ingredients); whether the article described the intervention as being theory-informed (and what theories informed the design); intervention format, length, and delivery; primary outcome(s) of interest; secondary outcome(s) of interest; proportion successful in comparison group(s) (as reported); proportion successful in intervention group (as reported); p-value and other relevant statistics regarding the main intervention effects on the primary outcome (with interpretation); whether the analytic approach was informed by theory; whether a mediation analysis was reported (if so, the analytic approach and variables tested as mediators); whether a moderation analysis was reported (if so, the analytic approach and variables tested as moderators); and any assumptions made about missingness. The authors created a table displaying all of the extracted data for easy comparison and examination. The lead author confirmed the accuracy of data extraction results.

#### Risk of bias assessment

Attrition, measurement (self-report vs biological, clinical, or some other form of objective measure), and differences between arms at baseline (e.g., with regard to substance use characteristics) were examined.

## Results

Ten studies met final inclusion criteria (total N=5410). All identified interventions sought to promote progress at one or multiple stages of the HIV care continuum, and several sought to address other outcomes as well (e.g., reduced drug use, reduced condomless sex). No interventions to promote PrEP continuum outcomes were identified.

### Study characteristics

Nine articles reflected studies wherein substance-using populations were intentionally targeted, while one article reflected a study with a population that incidentally used substances. Of the former, four studies involved use of at least one of several substances, including alcohol, opioids, stimulants, and/or marijuana, among others; three involved injection drug use only; one involved methamphetamine use only; and one involved alcohol use only. The latter study involved use of alcohol, injection drug use, and non-injection drug use. Seven studies utilized a two-arm RCT design, while two studies utilized a three-arm RCT design; one study utilized a four-arm RCT design. Geographic settings for studies included the United States (US; five), South Africa (one), Estonia (one), Vietnam (one), and Russia (one); one study spanned Indonesia, Vietnam, and Ukraine. Study quality was mixed, with attrition varying considerably across studies. Moreover, studies often used both biological testing (and/or clinical records) and self-report measures to assess outcomes (Table 1).

**Table 1.**
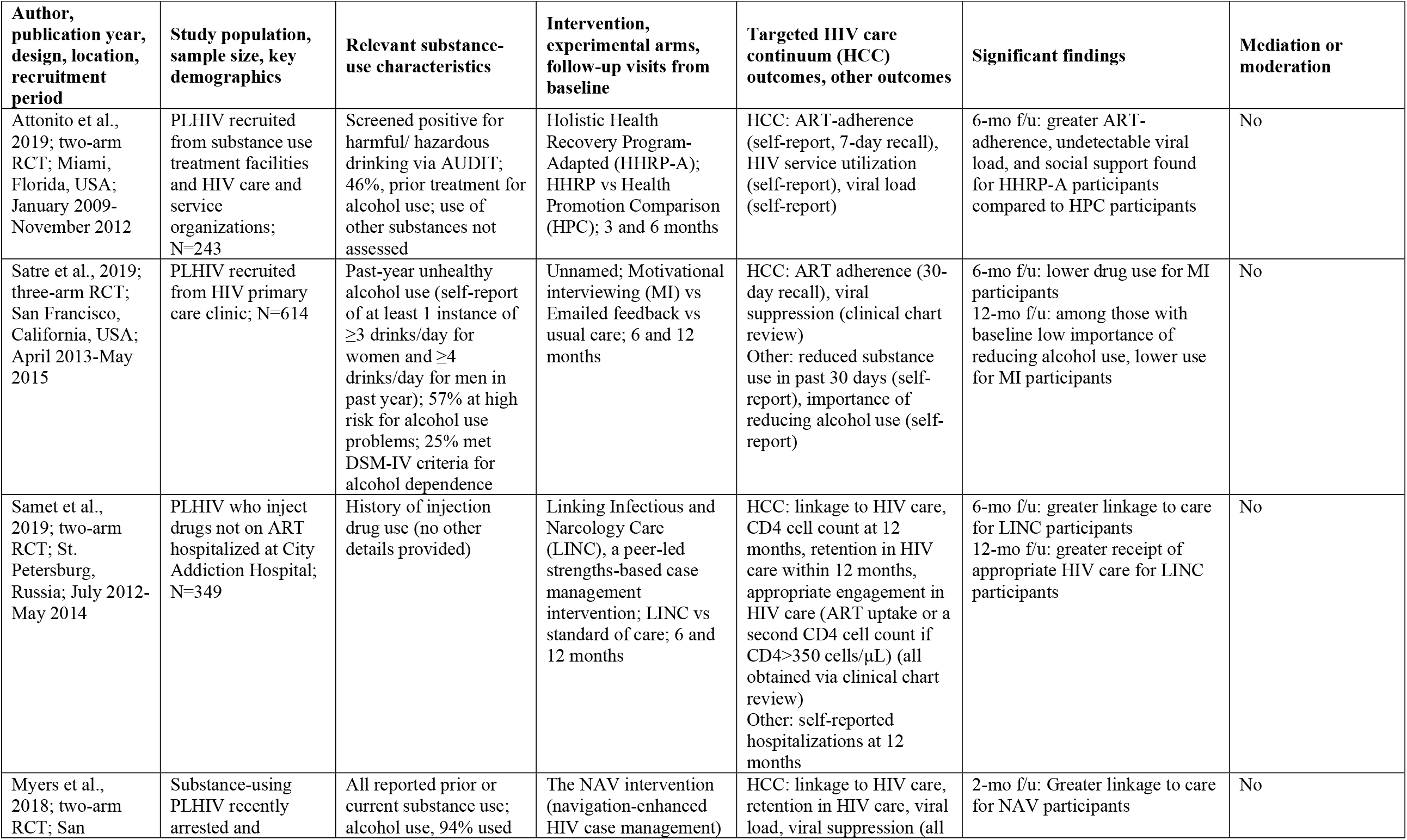
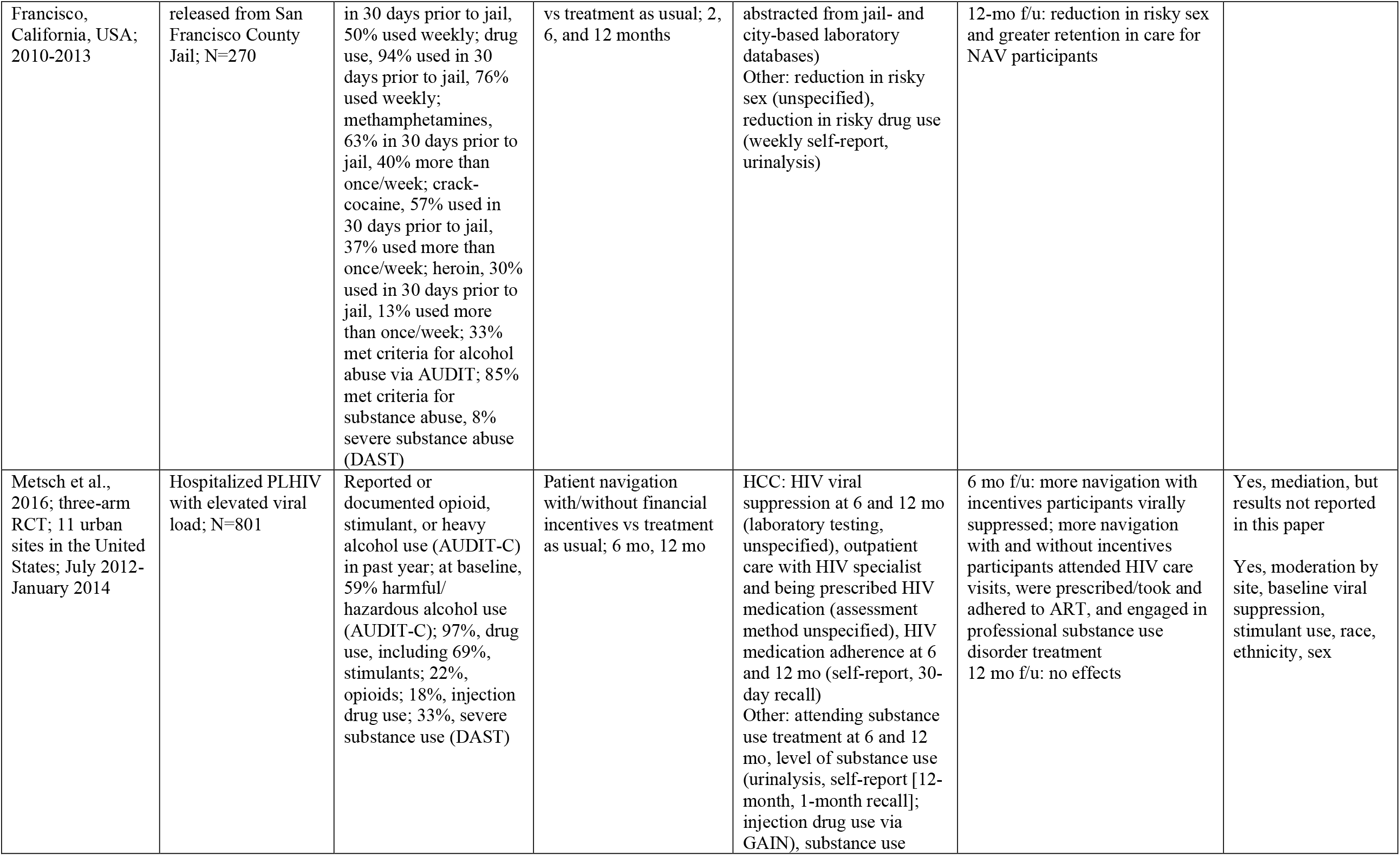
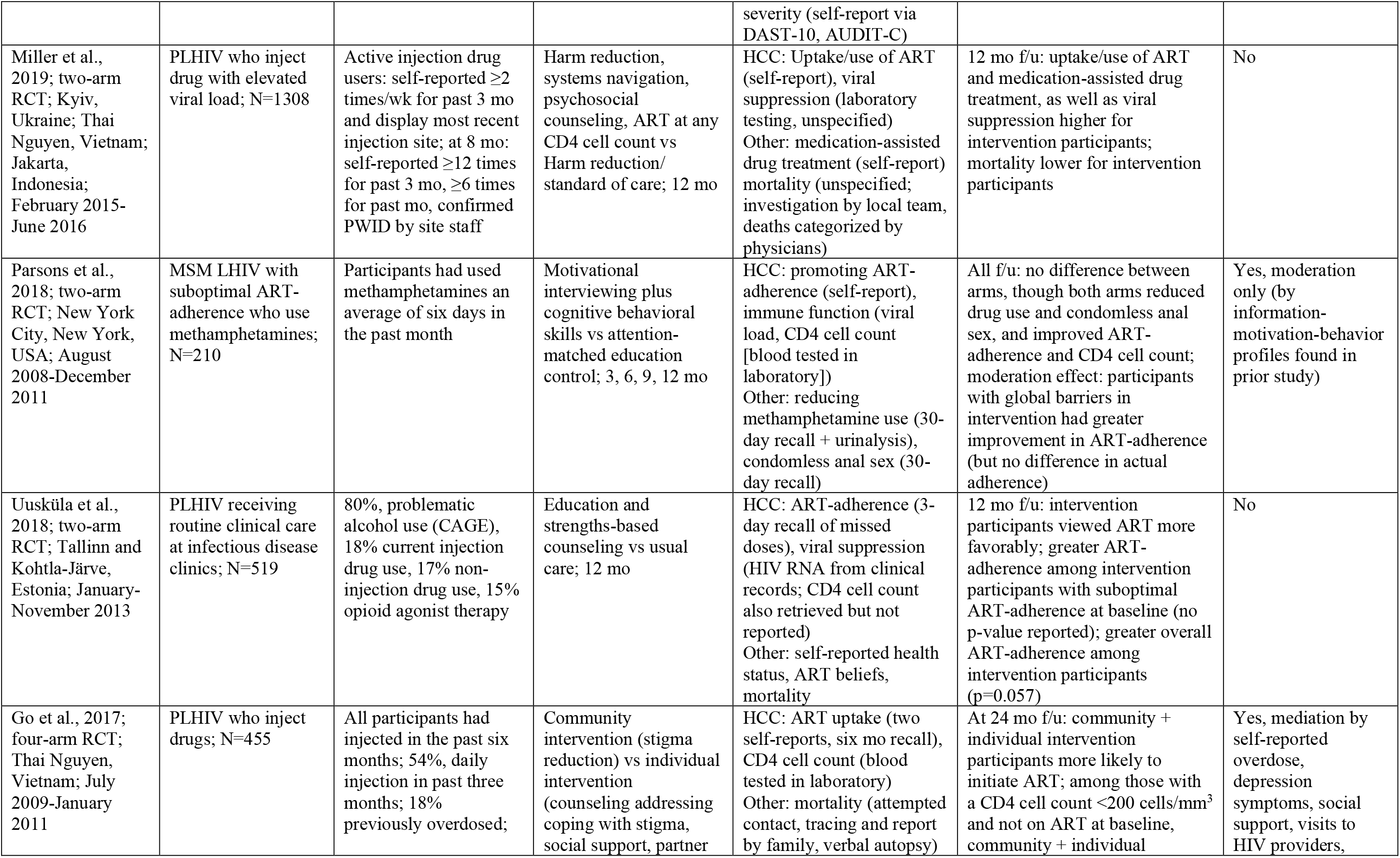
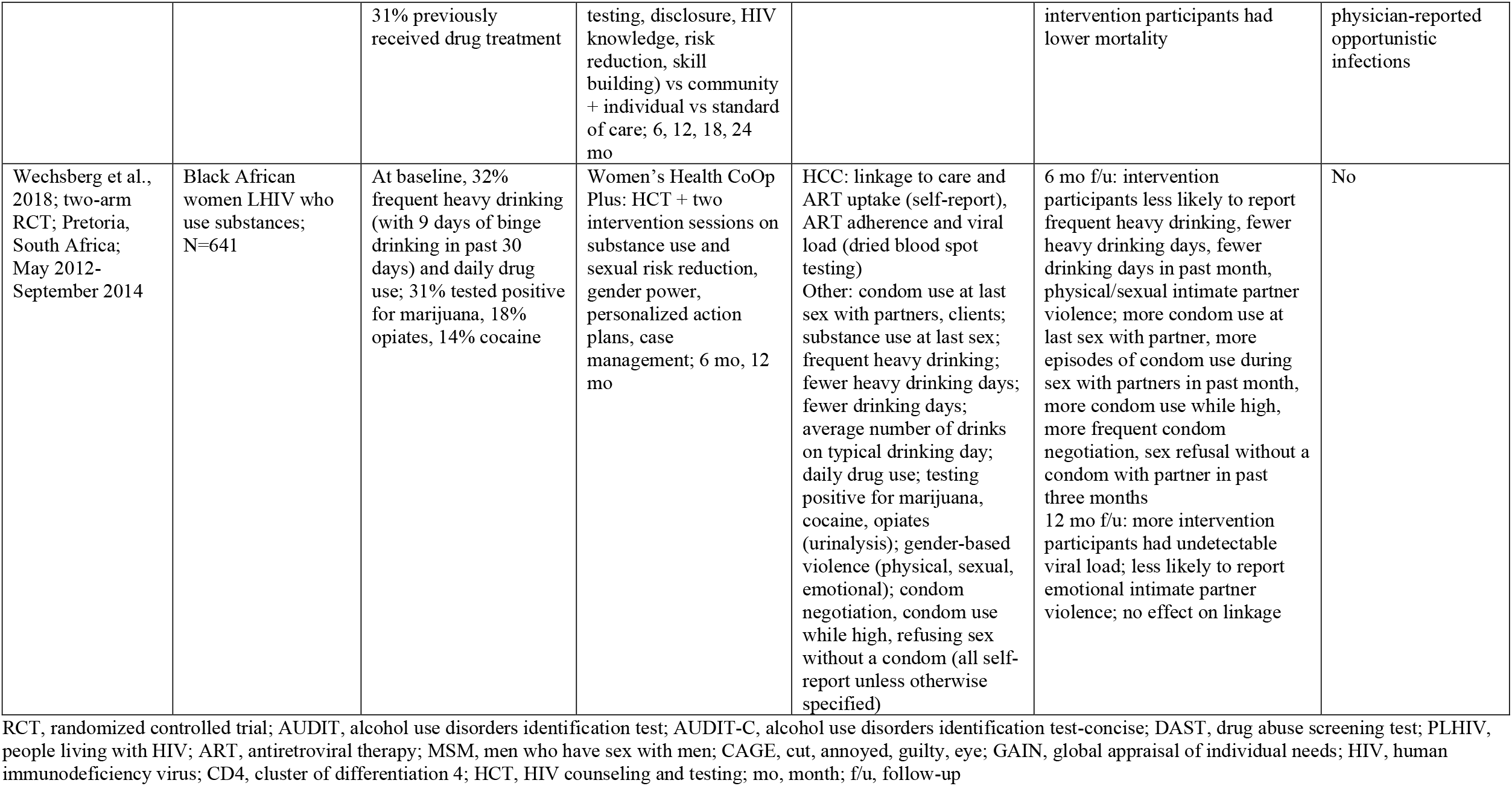
Description of included studies.

As with all systematic reviews, there are a range of characteristics (e.g., region, intervention type, drug type, outcome) by which HIV care continuum intervention studies with substance-using populations may be organized and discussed. In the results presented below, we center substance use: first, by whether or not substance-using populations were intentionally targeted, and second, by actual substance(s) used by those sampled. For each study, we briefly report the basic intervention components, each study’s sample, each sample’s substance use-related characteristics, and all significant intervention effects and other relevant findings, including those that do not directly pertain to the HIV care continuum. Additional information for each study is described in Table 1.

### Studies targeting substance-using populations

#### Multiple substances

Metsch et al. (2016) assessed the effect of a patient navigation intervention with and without financial incentives to promote retention in care, ART uptake and adherence, and viral suppression among hospitalized PLHIV with elevated viral loads and documented past-year opioid, stimulant, or heavy alcohol use in 11 urban hospitals across the US. At baseline, 59% of participants evidenced harmful/hazardous alcohol use, and 97% had documented stimulant, opioid, or other drug use; additionally, 18% had injected drugs in the past year, and 70% evidenced severe substance use.

##### HIV care continuum outcomes

At six-month follow-up, more navigation-with-incentives participants were virally suppressed, and more navigation-with-incentives participants and navigation-only participants had attended HIV care visits and had taken ART than control participants. There was no intervention effect on viral suppression at 12 months.

##### Other outcomes

At six-month follow-up, more navigation-with-incentives participants and navigation-only participants had engaged in professional substance use disorder treatment than control participants. There was no effect on other substance use outcomes at 12 months.

Myers and colleagues (2018) assessed the effect of a patient navigation-enhanced HIV case management intervention to promote linkage to HIV care, retention in HIV care, and viral suppression, as well as reduce risky sex and drug use behavior among PLHIV reporting prior or current substance use recently arrested and released from San Francisco County Jail. At baseline, 94% of participants reported alcohol use in the 30 days prior to jail, 50% of whom reported alcohol use more than weekly before jail; 94% reported drug use in the 30 days prior to jail, 76% of whom reported weekly drug use before jail. Methamphetamine use was the most reported drug used (63%; 40% used more than once per week), followed by crack-cocaine (57%; 37% used more than once per week) and heroin (30%; 13% used more than once per week). Thirty-three percent met criteria for alcohol abuse, and 85% met criteria for substance abuse, with 8% meeting criteria for severe substance abuse. Weekly drug use and using methamphetamines more than once per week in the 30 days prior to jail were both significantly higher in the intervention group relative to the control group.

##### HIV care continuum outcomes

Intervention participants were more likely to be linked to care within 30 days upon release and to be retained in care over the subsequent 12 months. Those who received substance dependence treatment in jail were more likely to be linked to care within 30 days upon release and to be retained in care over the subsequent 12 months. There was no effect on viral suppression.

##### Other outcomes

Intervention participants reported less risky sex at 12-month follow-up. There was no intervention effect on alcohol or drug use behaviors.

Satre et al. (2019) assessed the effect of an intervention (motivational interviewing vs emailed feedback vs usual care) to reduce unhealthy alcohol use, alcohol problems, and drug use, and promote ART-adherence and viral control among PLHIV with past-year unhealthy alcohol use recruited from an HIV primary care clinic in San Francisco. At baseline, roughly 57% of participants were at high risk for alcohol use problems, and roughly 25% met criteria for alcohol dependence (no differences across arms).

##### HIV care continuum outcomes

There were no effects on ART-adherence or viral control.

##### Other outcomes

There were declines in unhealthy alcohol use and alcohol problems within each arm but not between arms. At six-month follow-up, motivationally interviewed participants reported lower drug use/prescription drug misuse (excluding marijuana) than those in the other conditions. Among participants reporting baseline low importance of reducing alcohol use, those receiving motivational interviewing reported lower alcohol use at 12 months compared to those in the other conditions.

Wechsberg et al. (2019) assessed the effect of an intervention (risk reduction and gender power) to reduce drug and alcohol use, gender-based violence, and sexual risk, and to promote linkage to HIV care, ART uptake, viral suppression, and sexual negotiation among Black African women living with HIV who used at least one substance weekly for the past three months in Pretoria, South Africa. At baseline, 32% of participants reported frequent heavy drinking (with 9 days of binge drinking during the prior 30 days) and daily drug use; 31% tested positive for marijuana, 18% tested positive for opiates, and 14% tested positive for cocaine. Also at baseline, fewer intervention participants reported frequent heavy drinking (including days of binge drinking) than control participants, and fewer control participants reported daily drug use and tested positive for marijuana, opiates, and cocaine than intervention participants.

##### HIV care continuum outcomes

At 12-month follow-up, more intervention participants had undetectable viral load compared to control participants; there was no intervention effect on linkage to HIV care (among newly diagnosed, not-yet-linked participants) or ART uptake.

##### Other outcomes

At six-month follow-up, intervention participants were less likely to report frequent heavy drinking and reported fewer heavy drinking days and fewer drinking days in the past month. At six-month follow-up, intervention participants reported more condom use at last sex with partner, more episodes of condom use during sex with partners in the past month, more episodes of condom use while high, more frequent condom negotiation, and sex refusal without a condom with a partner in the past three months. At six-month follow-up, intervention participants were less likely to report physical or sexual intimate partner violence (and emotional intimate partner violence at 12-month follow-up).

#### Injection drug use

Go and colleagues (2017) assessed the effect of a multilevel stigma-reduction intervention on ART-uptake and survival among men who inject drugs (had injected in the past six months) in Thai Nguyen, Vietnam. At baseline, 54% reported daily injection drug use in the past three months, 18% reported prior overdosing, and 31% had previously received drug treatment (no differences across arms).

##### HIV-care continuum outcomes

Participants in the community-plus individual-level intervention were more likely to initiate ART than those in the standard-of-care condition.

##### Other outcomes

Among participants with a CD4 cell count <200 cells/mm^3^ (ART-eligible threshold at the time) and not on ART at baseline, those in the community-plus individual-level intervention had lower mortality than those in the standard-of-care condition.

Miller et al. (2018) assessed the effect of an integrated intervention (harm reduction, systems navigation, psychosocial counseling) to promote uptake and use of ART and medication-assisted drug treatment, and improve viral suppression among PLHIV who inject drugs with elevated HIV viral load in Kyiv, Ukraine; Thai Nguyen, Vietnam; and Jakarta, Indonesia.

##### HIV care continuum outcomes

At 12-month follow-up, uptake and use of ART, as well as viral suppression, were all higher for intervention participants relative to control participants.

##### Other outcomes

At 12-month follow-up, medication-assisted drug treatment was higher for intervention participants relative to control participants. Mortality was lower for intervention participants relative to control participants.

Samet and colleagues (2019) assessed the effect of a peer-led strengths-based case management intervention to promote linkage to care and CD4 cell count testing at 12 months among PLHIV who inject drugs hospitalized at the City Addiction Hospital in St. Petersburg, Russia. Other outcomes included appropriate HIV care (prescribed ART or a second CD4 cell count if CD4>350 cells/µL) and self-reported hospitalizations at 12 months.

##### HIV care continuum outcomes

At 6-month follow-up, more intervention participants had been linked to HIV care than control participants, and at 12-month follow-up, more intervention participants had received appropriate HIV care than control participants. There was no effect on CD4 cell count or retention in care.

##### Other outcomes

There was no effect on self-reported hospitalizations at 12 months.

#### Methamphetamines

Parsons et al. (2018) assessed the effect of a motivational interviewing plus cognitive behavioral therapy intervention on reducing methamphetamine use and condomless anal sex, and on promoting ART adherence and immune function (viral load, CD4 cell count) among cisgender sexual minority men living with HIV who use methamphetamines with suboptimal ART-adherence in New York City. At baseline, participants reported methamphetamine use an average of roughly six days in the past month (no difference between arms).

##### HIV care continuum outcomes

At 3-, 6-, 9-, and 12-month follow-ups, intervention and control participants evidenced increased ART adherence and CD4 cell count (and lower HIV viral load) compared to baseline, but there was no difference between arms. Moderation by Information-Motivation-Behavior class (identified in a prior study) emerged: among participants in the “global barriers” class, those in the intervention had a greater improvement in ART adherence than those in the control condition.

##### Other outcomes

At 3-, 6-, 9-, and 12-month follow-ups, intervention and control participants evidenced reduced substance use and condomless anal sex, but there was no difference between arms.

#### Alcohol

Attonito and colleagues (2020) assessed the effect of a holistic health intervention to improve ART adherence, service utilization, and HIV viral load among PLHIV reporting harmful/hazardous drinking recruited from substance abuse treatment facilities and HIV care and service organizations in Miami, Florida. Forty-six percent of participants reported prior treatment for alcohol use.

##### HIV care continuum outcomes

At six-month follow-up, intervention participants were more likely to report optimal ART-adherence (≥95%) and undetectable viral load. HIV service utilization improved for both arms, but there was no difference between arms.

##### Other outcomes

At six-month follow-up, intervention participants were more likely to report greater social support than control participants.

### Studies targeting populations with incidental substance use

#### Multiple substances

Uuskula and colleagues (2018) assessed the effect of an education and strengths-based counseling intervention intended to promote ART adherence and viral suppression among PLHIV receiving routine HIV clinical care from two infectious disease clinics in Tallinn and Kohtla-Järve, Estonia. Roughly 80% of participants screened for problematic alcohol use, 18% reported current injection drug use, 17% reported current non-injection drug use, and 15% reported being currently on opioid agonist therapy (with no differences between arms).

##### HIV care continuum outcomes

At 12-month follow-up, optimal ART-adherence (≥95%) was higher for those in the intervention group relative to the control group, which approached statistical significance (p=0.057). Among those with suboptimal ART-adherence at baseline, a higher proportion of intervention participants reported optimal adherence at 12 months relative to control participants (p-value not reported). There was no effect on viral load.

##### Other outcomes

At 12 months, intervention participants viewed ART more favorably than control participants, i.e., the differential in perceived ART need versus perceived ART concern was larger (p=0.046).

## Discussion

In this systematic review, we identified ten HIV care continuum intervention studies published since 2011 intentionally or incidentally targeting substance-using populations living with HIV. Populations using single or multiple substances, spanning alcohol, injection drugs, and non-injection drugs (including unspecified opiates, opioids such as heroin, stimulants such as methamphetamines and cocaine), were sampled in low-, middle-, and high-resource countries and ranged in size from N=210 to N=1308. A majority of interventions targeted individuals themselves (e.g., skill-building, reducing substance use), though some also targeted healthcare systems or communities in some way (e.g., patient navigation, case management, stigma mitigation). Interventions sought to address multiple HIV care continuum outcomes, including linkage to, receipt of, and retention/engagement in care; ART initiation (being prescribed ART, taking ART) and adherence; and viral suppression, with varying effectiveness. As no pattern emerged from the substance use-centered results (a likely reflection of the diversity of substances used across an already relatively low number of studies), we discuss the results by outcome.

It is unsurprising that viral load was the most commonly targeted HIV care continuum outcome (8/10 studies), as viral suppression – and ultimately viral undetectability – is the primary goal of HIV care continuum progression and motivated the transition to the TasP era (WHO, 2012). That greater viral load suppression was achieved at endpoints in only three interventions (with heavy alcohol-using, injection drug-using, and polysubstance-using samples) and at the midpoint only in one intervention (with a polysubstance-using sample) underscores the extent to which any substance use may substantially compromise HIV care efforts. However, the fact that intervention efficacy was demonstrated nonetheless shows promise for supporting viral load suppression among substance-using populations. The diversity of substances examined across the handful of efficacious studies (in addition to other factors) precludes the making of any overall deductions or conclusions about which substances may be most amenable to intervention to support viral load suppression. Further substance-specific research, including investigations of substances (e.g., stimulants) that may independently increase viral load (Carrico, 2011), is needed.

ART uptake/initiation and adherence were the second-most commonly targeted HIV care continuum outcomes (6/10 studies each), with greater ART initiation at endpoints in three interventions and at the midpoint in one intervention, and with greater ART adherence being achieved at endpoints in two interventions and at the midpoint in one intervention. ART uptake and adherence are of course necessary to achieve viral suppression and undetectability but remain vulnerable to the prioritization and effects of substance use (Kuchinad et al., 2016). As alcohol was the primary substance used by the samples in the studies in which intervention effects were found, ART adherence in the context of heavy alcohol use may be particularly amenable to intervention but requires further investigation. ART adherence in the context of other substance use, as well as ART adherence as a mediator between intervention and viral load suppression and undetectability in trials are also areas for future research.

Retention in care was a less commonly targeted outcome (4/10 studies: two explicitly identified retention in care as an intervention target, two identified HIV service utilization and attending HIV care visits as intervention targets). Greater retention in care was achieved at the endpoint of one intervention and at the midpoint of another. Retention in care may have been minimally examined because it may be more commonly viewed as the responsibility of healthcare systems rather than healthcare patients. However, retention in care remains an important stage of the HIV care continuum leading to viral suppression, and clear disparities in retention in care have been demonstrated for people who inject drugs (or have a history of such) relative to those who do not or have not injected drugs (Anderson et al., 2020). Interventions targeting retention in care among people who inject drugs or have a history of injection drug use may be warranted.

Linkage to care was also less commonly targeted (3/10 studies), possibly due to the fact that universal test-and-treat strategies were being or beginning to be implemented during our pre-specified time period for article inclusion (2011 onward), or due to linkage to care being framed similarly to retention in care (healthcare system vs individual responsibility) (Cohen, McCauley, & Gamble, 2012; Hontelez et al., 2013; Smith, Powers, Kashuba, & Cohen, 2011; Wagner & Blower, 2012). However, because no main intervention effects on linkage to care were retained at any study’s endpoints, linkage to care may be an appropriate intervention target for future trials. Relatedly, none of the identified interventions targeted HIV diagnosis or status awareness as a precursor to linkage to care and entering into the HIV care continuum. Substance use can lead to delays and even intentional avoidance of HIV testing, as well as delayed linkage to HIV care among PLHIV (Kuchinad et al., 2016). Like linkage to care, HIV testing and status awareness may be inadvertently neglected in intervention efforts with substance-using populations living with HIV and may need to be reconsidered in future intervention development and testing endeavors.

Though we were unable to draw any conclusions regarding specific substances’ amenability to intervention, we did identify an important similarity among interventions shown to be efficacious at their endpoints. In addition to targeting individual-level factors, these interventions expanded beyond individuals and targeted broader factors – such as interpersonal-level (social support), healthcare system-level (patient navigation, case management), or community-level factors (stigma mitigation) – to varying extents. Though substance use is an individual behavior and some HIV care continuum stages involve individual behaviors (e.g., ART adherence), such behaviors are not enacted in a vacuum. A range of factors across socioecological levels operates to shape substance use, as well as HIV prevention and care efforts (Amaro, Sanchez, Bautista, & Cox, 2021; Baral, Logie, Grosso, Wirtz, & Beyrer, 2013; Lacombe-Duncan et al., 2019; Sullivan et al., 2021). Interventions targeting multiple, especially higher-order socioecological levels may be more effective for supporting substance-using populations.

Mediation and/or moderation analyses were infrequently conducted in the identified studies, potentially masking additional findings that may prove to be statistically, clinically, or behaviorally significant. Revisiting trials whose outcome analyses were restricted to main effects averaged over all participants and reanalyzing data from those trials (whether originally efficacious or not) by including mediators and/or moderators in analytic models may reveal significant findings undetected initially, including new intervention to outcome pathways or previously unidentified subgroups for whom the intervention demonstrated greater or lesser efficacy (Pitpitan et al., 2021). Notably, several of the studies in this review did report significant intervention effects on non-HIV care continuum outcomes (e.g., reduced substance use, condomless sex, gender-based violence; increased social support). These other outcomes may act as mediators or moderators of the intervention effect on HIV care continuum outcomes and should be explored in future research, whether through secondary analyses of data from these completed trials or through planned analyses of new intervention trials.

We did not identify any PrEP-related studies that met our inclusion criteria in our search. However, screening and examination of PrEP-related articles for inclusion in this review suggested that there is growing intervention attention to promote engagement in the PrEP care cascade among people who use drugs, yielding four protocol papers for intervention studies that are currently underway for this population (Arrington-Sanders et al., 2020; Shrestha, Altice, Sibilio, Ssenyonjo, & Copenhaver, 2019; Starks et al., 2019; Wechsberg et al., 2020). Moreover, a systematic review of the state of the PrEP care cascade among people who inject drugs (Mistler et al., 2021) described a feasibility and acceptability study for an intervention to increase PrEP adherence among people who inject drugs (Shrestha, Altice, Karki, & Copenhaver, 2018). Prompt and rigorous evaluation of these interventions will be useful for understanding how to support people who use drugs and alcohol progress across the PrEP care cascade.

### Limitations

Since our search included terms related to drug, alcohol, or substance use, we may have failed to locate studies in which populations with incidental substance use were sampled, as such studies may not have been indexed using those terms at the time of publication or may have reflected interventions that did not target substance-using populations. Second, several factors prevented easy between-study comparisons and limited generalizability of findings: the diversity of substances and drug use behaviors examined across studies, the variations in substance use measurement (for studies that did involve the same substance or behavior), the range of countries (low and middle income versus high income) and contexts (urban cities; hospitals, jails) where studies were conducted, and the different HIV treatment policies in place at the times studies were conducted. Third, several studies examined self-reported rather than objectively or biologically measured outcomes, potentially resulting in reporting bias. Relatedly, comparatively low attrition in some studies, as well as differences in substance use characteristics between study arms at baseline, may have biased findings. Finally, we focused only on intervention outcomes published since 2011. A review inclusive of studies published prior to the TasP era may have yielded a different picture of the landscape of HIV care continuum outcomes among people who use substances.

## Conclusions

Globally, substance use remains both a prevalent health behavior and a barrier to HIV care continuum progression among PLHIV. Several interventions with people who use substances in the TasP era have been conducted, with roughly half demonstrating some level of efficacy in achieving targeted HIV care continuum outcomes. However, much about intervention effects remains unknown, given the low number of studies identified here, as well as the diversity of substances examined and the nuanced effects (direct and indirect) that various substances may have on HIV-related outcomes. Additional intervention research specific to the use of certain substances across diverse samples, as well as research involving the re-examination of data from previous trials to tease out mediated and moderated intervention effects, can illuminate more clearly how to support PLHIV who use substances.

## Supporting information

prisma checklist

prisma checklist for abstracts

coi pitpitan

coi baker

coi bejarano

coi wiginton

## Data Availability

Our specific collection of data that we extracted from the studies are not currently available; however, as this is a systematic review, all data included in the review are available in the original articles from which the data were extracted.

